# GABAergic regulation of action-outcome priors in conditions associated with frontotemporal degeneration

**DOI:** 10.1101/2025.10.22.25338552

**Authors:** Rebecca S. Williams, Michelle Naessens, Emily G Todd, Robert Durcan, David J Whiteside, Juliette Lanskey, Amirhossein Jafarian, Karl Friston, Laura E. Hughes, James B. Rowe

## Abstract

**Rationale:** Apathy is a common symptom in many neurological and psychiatric conditions, associated with poor prognosis and increased caregiver burden. There are currently no proven treatments, in part due to a lack of mechanistic understanding. We have proposed a novel framework for apathy, based on the failure of active inference due to imprecise priors on the outcome of actions. The loss of precision on action outcomes causes apathy by reducing the expected difference between the state of the world following action versus non-action. Here we test the hypothesis that the loss of prior precision on action outcomes is reversible and mediated neuronally by GABAergic gain on the superficial pyramidal neurons in a prefrontal-motor decision-making hierarchy. We test this in a healthy cohort and people with two syndromes associated with frontotemporal lobar degeneration.

**Methods:** Twenty healthy controls, twenty people with behavioural variant frontotemporal dementia (bvFTD) and twenty people with progressive supranuclear palsy (PSP) took part in a randomised placebo-controlled double-blind trial using zolpidem, an established GABAA agonist. This study was registered with ISRCTN registry (ISRCTN10616794). We use the ‘Goal Prior Assay’ task and dynamic causal modelling of MEG resting-state data to explore the cognitive and neural concomitants of prior precision, and the effect of GABAergic regulation on both prior precision and superficial pyramidal gain. Apathy was primarily assessed with the Apathy Evaluation Scale (Self and Carer). Principal analyses were conducted using Bayesian statistics, supplemented by classical frequentist tests.

**Results:** Forty-three participants (20 controls and 23 patients) were included in the final analysis. We found strong evidence for a difference in measures of apathy (B>1000, p<0.001) and prior precision (B=20.4, p<0.01) between healthy controls and people with bvFTD and PSP. This difference in prior precision was not found in the drug condition (B=0.86, p=0.11). There was strong evidence of a correlation between apathy and prior precision across groups (B>100, p<0.001). Dynamic causal modelling of MEG resting-state data confirmed reductions in gain on the prefrontal superficial pyramidal neurons in patients. The prefrontal superficial pyramidal gain was partially restored on zolpidem and linked to participants’ prior precision on their action outcomes.

**Conclusion:** We confirm that apathy in conditions associated with frontotemporal lobar degeneration is underwritten by a reduction in prior precision on action outcomes, mediated by reduced synaptic gain of prefrontal superficial pyramidal neurons. GABAergic regulation using zolpidem partially restores prior precision by acting on this gain and reinstating neuronal message-passing within the prefrontal-motor decision-making hierarchy.

## Introduction

Apathy is a common symptom in many neurological and psychiatric disorders, including syndromes associated with frontotemporal lobar degeneration^1^ (FTLD). It is defined as a reduction in goal-directed action^2^, and predicts both functional independence^3^ and mortality^4^ over and above age or cognitive ability. Apathy is positively associated with caregiver burden^5,6^ and is a priority target symptom for treatment in dementia by both clinicians and carers^7,8^.

While preclinical models of apathy have emphasised the role of reward and dopaminergic depletion, apathy can also be considered as the result of a failure of active inference, resulting from the loss of prior precision on action outcomes^9^. In the ‘Bayesian Brain^10–12^, prior beliefs are key to learning accurate models of the environment, including action outcomes. Priors are combined with sensory evidence to update beliefs about the world, allowing us to predict and interact with our surroundings. When there is a discrepancy between predicted outcomes and sensory observations, beliefs may be updated passively (i.e., changing one’s perception or understanding of the situation), or one may actively shape the environment, so that it better matches with prior expectations (active inference)^13^. Prior precision, characterised as the certainty or confidence afforded a prior belief, is a key factor in active inference. When prior precision on action outcomes is very low, the difference between the expected consequences of acting *versus* not-acting disappears. For example, if your beliefs about how much warmer you will be with a jacket on are imprecise, you are less likely to take the action of putting a jacket on to keep warm. This prior precision is a distinct property from the magnitude of the expected reward. Apathy can therefore arise as a failure of active inference due to imprecise priors on action outcomes, as there is an implicit uncertainty about how actions will bring the world closer to a goal state. This hypothesis is supported by the association between apathy and prior precision in healthy individuals and in people with Parkinson’s disease^9,14,15^.

There are currently no proven pharmacological treatments for apathy^16^. Selective serotonin reuptake inhibitors are commonly used to treat apathy in FTLD^8^ despite mixed evidence of efficacy^17–19^ and concerns of worsening amotivational syndromes^20^. Similarly, selective dopamine agonists and levodopa in isolation do not reliably treat clinical apathy^16,21–23^. However, compounds with joint noradrenergic and dopaminergic action like methylphenidate have shown some positive effects^24–26^, including in neurological conditions such as Alzheimer’s disease^24^. Noradrenaline has also been linked to the related processes of effort processing^27^, learning under uncertainty^28^, and prior precision^29^. Such noradrenergic effects on prior precision may be direct, or indirect, through interactions with gamma-aminobutyric acid (GABA)^30–34^.

GABAergic deficits are well-established in FTLD-associated conditions, with evidence of reduced pyramidal neurons^35^ and GABA-receptors^36,37^ in the frontal and temporal lobes. A GABA deficit has also been confirmed *in vivo* in behavioural variant frontotemporal dementia (bvFTD) and progressive supranuclear palsy (PSP) using magnetic resonance spectroscopy^38^. The consequence of these GABA deficits has been further characterised using dynamic causal models of human cortical physiology^39^. GABA is critical in maintaining the dynamic modulation of information processing in the frontal cortex^40^, and its loss results in effective disconnection in networks for goal-directed decision-making. In these networks, modulation of synaptic gain on superficial pyramidal cells by GABA is proposed to encode or mediate prior precision^40^. If confirmed, this hypothesis would open new GABAergic strategies to normalise prior precision, and in turn treat apathy.

We test this hypothesis using zolpidem, a GABA_A_ receptor agonist (licensed as a sedative-hypnotic). Zolpidem modulates GABA levels and glucose metabolism in human prefrontal cortex^41^ and has been used off-label for the treatment of symptoms in disorders of movement^42^ and consciousness^43^, with small scale studies of severe amotivational states reporting benefits^41,44,45^. Preclinical studies also demonstrate potential anti-apathy mechanisms^46^, with the potential for broader clinical impacts highlighted in conditions like PSP^47^. We therefore selected zolpidem to test the hypothesis that GABAergic modulation may improve prior precision in people at risk of a GABAergic deficit as a result of bvFTD or PSP.

We undertook a randomised placebo-controlled double-blind trial to test the neural and computational mechanisms underpinning apathy. Specifically, we hypothesised that zolpidem would improve prior precision in individuals with bvFTD and PSP, but not in healthy controls, by restoring GABAergic neuromodulatory mechanisms in the prefrontal cortex. We investigate these neural mechanisms using dynamic causal modelling of resting-state MEG data, with a focus on the gain of superficial pyramidal neurons in the prefrontal cortex. This mechanistic study is not a Clinical Trial of an Investigational Medicinal Product (i.e. not a CTIMP), but confirmation of the hypothesis would indicate the potential for future CTIMPS to evaluate GABAergic drugs to reduce apathy.

## Materials and methods

### Participants

Twenty people with probable behavioural variant frontotemporal dementia (bvFTD^48^), 20 people with probable progressive supranuclear palsy (PSP - Richardson syndrome^49^) and 20 healthy controls were recruited. Recruitment for this stage of the study took place from March 2022 to January 2025. The controls were recruited from either the volunteer panel of the MRC Cognition and Brain Sciences Unit or the Join Dementia Research register. Healthy volunteers were included if they were right-handed, aged between 50 and 75 years old, and had normal or corrected-to-normal vision and hearing. Volunteers with probable bvFTD or PSP were recruited from the NHS dementia clinics. Exclusion criteria in both groups included those currently taking GABAergic medications, and individuals with a history of brain injuries. Normal clinical care continued through participation in the study with no change in concomitant medications.

Diagnoses were made by a consultant neurologist at a multidisciplinary clinic, based on international consensus criterion. Individuals with other types of dementia, or PSP phenotypes, were not included. The study was registered with ISRCTN registry (ISRCTN10616794; https://www.isrctn.com/ISRCTN10616794) and the protocol was exempted from clinical trials status by the Medicines and Healthcare Products Regulatory Agency, UK, as a mechanistic study with non-clinical primary outcomes. Participants gave written informed consent according to the Declaration of Helsinki, and the study protocol was approved by the local Research Ethics Committee. The duration of the study days, disease severity, behavioural and technical factors meant that approximately half of patients were able to complete the task with sufficient data and quality for analysis.

There are clinical and neurological distinctions between bvFTD, and PSP. However, there is also significant clinical overlap between the two, including apathy, prefrontal neurophysiological deficits and GABAergic deficits^39^. We therefore collapsed across diagnostic groups for principal analyses and separated them for secondary tests.

### Procedure

Participants completed three testing sessions. The first two sessions constituted a randomized double-blind placebo-controlled crossover design and were conducted approximately two weeks apart, at the same time of day. Participants were given a single dose 5mg oral tablet of zolpidem or matched placebo. The randomisation order was permuted in sequential blocks of six participants and known only to the dispensing pharmacist. The Clinical Research Facility team (independent of the study team) administered the tablets on each visit at Addenbrookes Hospital, Cambridge, UK. Venus blood samples were taken approximately 90 minutes after drug administration, immediately before the MEG recording, close in time to the estimated peak plasma levels and central nervous system penetration. MEG recordings took place at the MRC Cognition & Brain Sciences Unit, University of Cambridge, UK. At the start of each session, participants completed a 5-minute resting-state MEG recording with eyes closed. At the end of the session, participants completed the ‘Goal Prior Assay’ paradigm^14^ (detailed below) out-of-scanner. The third testing session included 7-Tesla MRI — for source reconstruction of the MEG data — at the Wolfson Brain Imaging Centre, University of Cambridge, UK.

### Materials

The primary behavioural task in this study was the ‘Goal Prior Assay’ task, developed by Hezemans et al. (2020)^14^. This task was designed to infer the precision of prior beliefs and its influence on the perception of action outcomes. The aim of the task is for participants to land a virtual ball on a target by applying sufficient power to a force sensor. In a pseudo-random subset of trials the ball disappears before reaching its end position, and participants are asked to estimate where they believe the ball’s final position to be (see *figure 1*).

**Figure 1.**
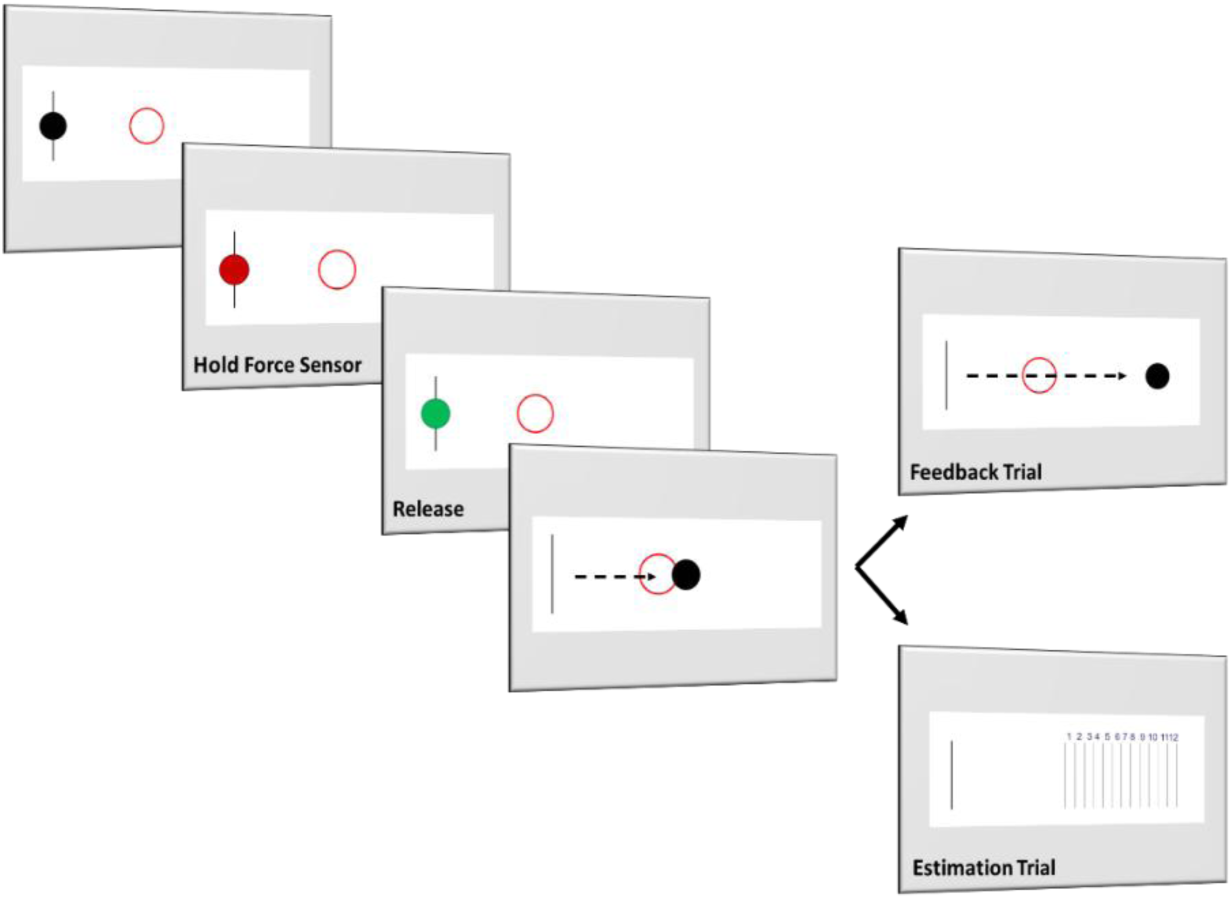
The ‘Goal Prior Assay’ task originated by Hezemans et al (2020). When the ball appears, participants are asked to press a force sensor with sufficient force, sustained for 3s, for it to reach the target under learned stable kinematics. On pressing the sensor, the ball turns red. After 3 seconds, the ball turns green, and participants release the force sensor. The virtual ball then travels across the screen, with initial velocity scaled to the average force applied to the sensor, subject to constant deceleration. In non-catch trials, the ball is seen to stop. In pseudo-random catch trials, the ball disappears early in its trajectory and participants are asked to estimate the final position of the ball using an estimation scale that appears on screen.

The task consisted of 120 trials, including 40 estimation trials, with 15 additional practise trials, which are excluded from analysis. The trials are split into 4 blocks, randomised in their presentation. 2 blocks are considered ‘high effort’ (i.e. the target is farther away, requiring high force) and 2 blocks ‘low effort’ (i.e. the target is closer, requiring low force).

### MEG Data Acquisition

MEG data were acquired at 1 kHz in a magnetically shielded room using the 306-channel MEGIN TRIUX neuro system which includes 204 planar gradiometers and 102 magnetometers. Eye movements were measured using pairs of vertical and horizontal EOG electrodes. Prior to data acquisition, a 3D digitizer (Fastrak Polyhemis Inc.) was used to record (1) three fiducial points, one on the nasion and one each on the left/right pre-auricular points, (2) the location of five HPI coils, and (3) ∼300 ‘head points’ across the scalp.

The preprocessing pipeline for our resting state data is based on Vaghari et al (2022; https://github.com/delshadv/BioFIND-data-paper). MEG preprocessing was conducted in MATLAB v2018a using the Statistical Parametric Mapping (SPM12) toolbox. Raw MEG data were preprocessed using Maxfilter software (version 2.2.14, Neuro-TRIUX), including standard signal-space separation, movement compensation and translation to default space. The data were segmented into epochs of 1 second duration and downsampled to 500Hz, and filtered using a 0.5-150Hz bandpass filter followed by 48-52Hz notch filter. Artefact rejection used the OHBA Software Library. Finally, the cleaned data was manually coregistered to a 7T scan (N=28) using the fiducials digitised at MEG acquisition. Where a 7T MRI scan was not available, a volumetric 3T MRI scan was instead used for coregistration (N=10). Where neither was available, we used a symmetrical 7T template (https://www.templateflow.org/^51^). One participant was unable to tolerate either an MEG or MRI scan.

7T MRI used a Siemens MAGNETOM Terra scanner with a 32-channel head receiver and single channel transmit head coil (Nova Medical) at the Wolfson Brain Imaging Centre, University of Cambridge. A high resolution isotropic whole brain Magnetization Prepared 2 Rapid Acquisition Gradient Echoes (MP2RAGE) sequence (repetition time=4300 ms, echo time=1.99 ms, resolution=99 ms, bandwidth=250 Hz/px, voxel size=0.75 mm3, field of view= 240 × 240 × 157mm, acceleration factor (A ≫ P) =3, flip-angle=5/6° and inversion times=840/2370 ms) was acquired for the purposes of MEG coregistration. 3T MRI used a Siemens PRISMA scanner, with a 32-channel head receiver and single channel transmit head coil (Nova Medical) with T1-weighted Magnetization Prepared Rapid Acquisition Gradient Echoes (MPRAGE; TR=2000ms, TE=2.93ms, TI=850ms, FA=8°, 208 slices, 1.1mm isotropic voxels).

### Modelling and Statistical Analysis

Principal analyses were conducted using Bayesian statistics. For completeness, frequentist statistics are also reported but note that they are not interchangeable tests as the nature of the inferences supported is different. The Bayes Factor (B) is interpreted in keeping with convention: standard evidentiary thresholds are used for moderate (>3), strong (>10) or very strong (>100)^52,53^ evidence in favour of the alternate hypothesis, or 1/3, 1/10 and 1/100 for corresponding strength of evidence for the null hypothesis. In the case of frequentist statistics, results are considered significant if p<0.05.

Participants with relevant data missing were excluded at the level of individual analyses. Analyses comparing the demographic and clinical characteristics of responders and non-responders to the ‘Goal Prior Assay Task’ are reported in Supplementary Material 1.

Code for this data analysis pipeline is available at https://github.com/BeccaSue99/prior-GABA.

### Behavioural

Behavioural analyses were conducted with custom scripts using R (v4.3.1) with RStudio (2023). Our primary hypothesis is that individuals with bvFTD and PSP are (a) more apathetic and (b) have more imprecise priors relative to healthy controls. This was analysed using two Bayesian pseudo-t-tests, comparing apathy scores and prior precision between patients and controls. Apathy was measured using the Apathy Evaluation Scale (AES; Self and Carer)^54^ and the motivation subscale of the Cambridge Questionnaire for Apathy and Impulsivity Traits (CamQUAIT-M)^55^. Prior precision in this task was operationalised as the best-fit regression slope between performance and estimation error on the ‘Goal Prior Assay’ task. Performance error was the difference between the target and final ball position, whereas estimation error was the difference between the estimate and final ball position. Participants with greater prior precision should be biased towards the location of the target (i.e. their action outcome prior), meaning a negative association between performance and estimation error. The gradient of the regression slope can therefore be used as a proxy of prior precision.

We also hypothesised that (a) individuals with FTLD-associated conditions on zolpidem would have more precise priors than those on placebo, but (b) this would not be the case for healthy controls. Given the established GABA deficits in FTLD^56^, we hypothesise that zolpidem may succeed in increasing GABA levels and improve prior precision through augmenting cortical gain on the prefrontal superficial pyramidal neurons. However, as healthy controls have normal GABA levels, we hypothesised that zolpidem may lead to over-dosing of GABA resulting in no improvement, and potentially worsening of, prior precision. We tested these hypotheses using Bayesian pseudo t-tests. We analyse the significance of drug-group interaction by comparing evidence for linear regression models with and without an interaction term.

Our co-primary hypothesis is that there is a positive correlation between prior precision and apathy. This is a replication of previous work by Hezemans et al. (2020), implemented in a new diagnostic group and assessed using a Bayesian correlation.

### Dynamic causal Modelling

We implemented dynamic causal modelling (DCM) of cross spectral densities to analyse resting-state data of participants in MEG. DCM is a generative modelling procedure designed to characterise the synaptic connectivity of neuronal networks using neural mass models and Bayesian modelling procedures (i.e., Variational Laplace)^57–59^. DCM operates by identifying a biologically-plausible neuronal model which is inverted and fitted to the dataset by varying key parameters, such as intrinsic connectivity between neuronal populations within a cortical source, and extrinsic connectivity between sources. We predicted a reduction in gain of the superficial pyramidal neurons in the prefrontal cortex and a reduction in the top-down extrinsic connectivity needed for integration of prior beliefs with sensory evidence.

Based on prior literature^60^ and previous modelling of the ‘Goal Prior Assay’ task in scanner^9^ we selected three sources of interest in our model: right inferior frontal gyrus (pars orbitalis) [56; 26; -12], left premotor cortex [-14; 16; 58], and left motor cortex [-37; -25; 64].

We investigated the role of diagnostic group, drug condition and prior precision in modulating estimates of connectivity within and between these three sources using Parametric Empirical Bayes (PEB)^61^.

## Results

To ensure effective estimates of prior precision, participants were included in analysis if they completed two or more of the four testing blocks (with catch trials > 20). This led to the inclusion of 20 controls and 17 patients in the placebo condition, and 19 controls and 21 patients in the zolpidem condition. Differences in demographics and clinical scores between patient responders and non-responders is available in Supplementary Material 1. 19 controls and 15 patients had complete data from both visits. There were no unexpected adverse reactions.

Demographics of the participants with complete data from one or both sessions is available in Table 1. There was no evidence of a difference in age between controls and patients (*B=0.36, p=0.53*), though there was strong evidence for a difference in years of education (*B=29.3, p<0.01*) with patients having less education than controls. There was also very strong evidence of a difference in ACE-R and CBI-R between controls and patients, with patients being significantly more impaired (*B>100, p<0.001*).

**Table 1.** Demographic and diagnostic data for participants with usable task data in one or both conditions. Values presented are the mean and standard deviation of available data for each metric with comparisons analysed using Bayesian and frequentist t-tests, except gender differences which was analysed using Bayesian contingency tables and a chi-squared test.

|  | Controls (N=20) | Patients (N=23) | Comparison |
| --- | --- | --- | --- |
| Age (SD) | 66.6 (7.04) | 68.05 (7.82) | B = 0.36 (p=0.53) |
| Gender (M: F) | 11:9 | 16:6 | B = 1.03 (p=0.38) |
| Education, years (SD) | 16.1 (2.88) | 12.74 (3.29) | B = 29.3 (p<0.01) |
| bvFTD: PSP | - | 11:12 | - |
| Mean ACE-R (SD) | 96.35 (3.25) | 83.05 (11.1) | B>100 (p<0.01) |
| MMSE (SD) | 29.53 (0.71) | 26.43 (3.72) | B = 19.47 (p<0.01) |
| CBI-R (SD) | 4.13 (4.77) | 69.3 (44.56) | B>100 (p<0.01) |
| CamQUAIT-M | 3.47 (3.52) | 14.85 (6.54) | B>100 (p<0.01) |
| AES-Self | 25.6 (4.61) | 37.13 (9.79) | B>100 (p<0.01) |
| AES-Carer | 24.2 (5.78) | 46.7 (12.67) | B>100 (p<0.01) |

### Behavioural

There was very strong evidence of a difference in apathy scores between controls and patients for the AES-Self (*B>1000, p<0.001, d=1.64*), AES-Carer (*B>1000, p<0.001, d=2.42*) and motivation subscale of the CamQUAIT (*B>1000, p<0.001, d=2.26*) such that patients scored significantly higher than controls.

The median prior precision in controls was *0.49* in the placebo condition and *0.53* in the drug condition. The median prior precision in patients was *0.12* in the placebo condition and *0.32* in the drug condition (see *figure 2*). There was no evidence for a difference in prior precision between patients with a diagnosis of bvFTD or PSP (*B=0.45, p=0.64, d=0.21*), and so groups remained combined for subsequent principal analyses.

**Figure 2.**
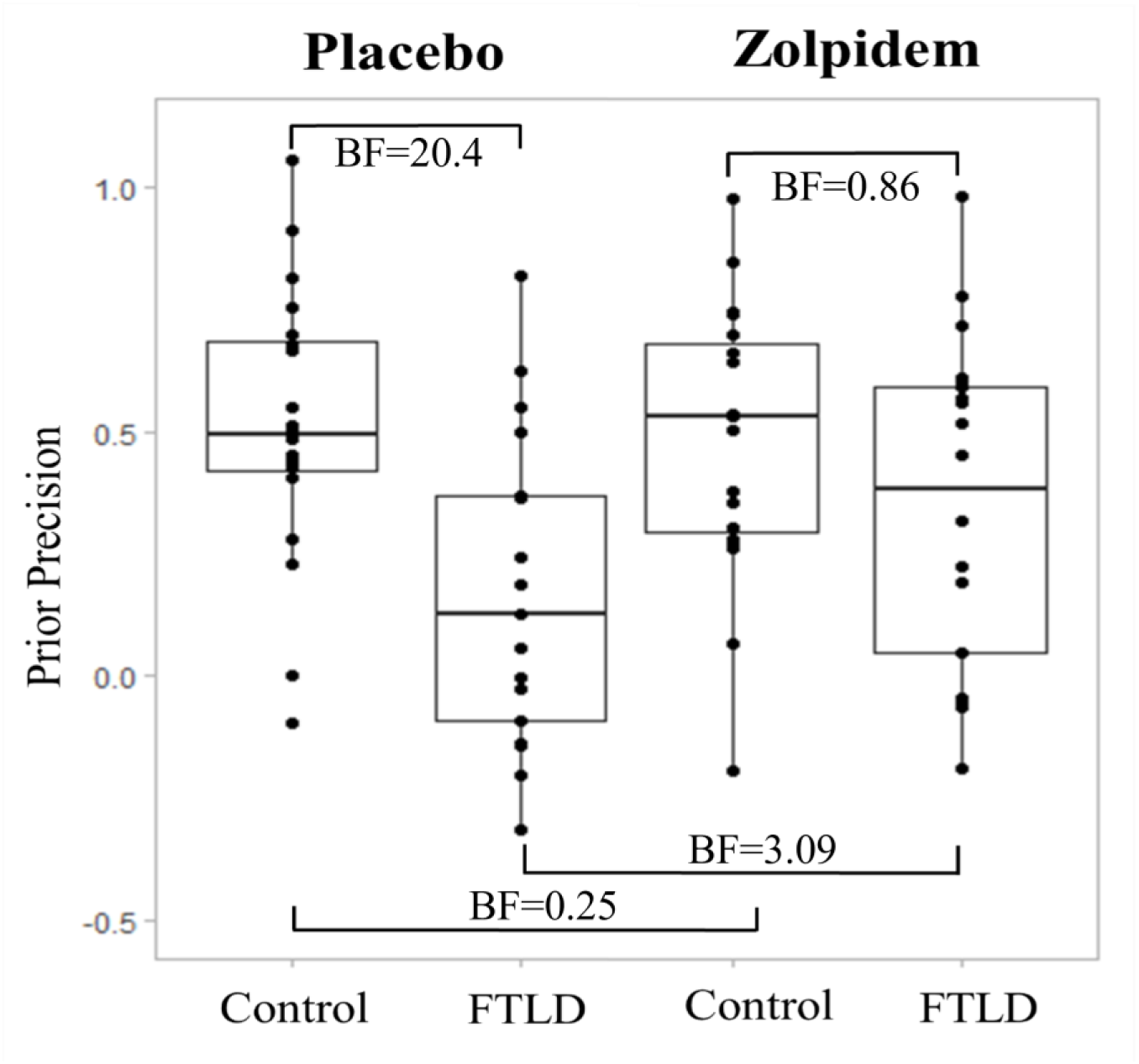
Prior precision in controls and patients with FTLD-associated disorders, in placebo and zolpidem conditions. Higher values indicate higher levels of prior precision.

The critical result is that there was evidence for an improvement in prior precision in the patient group when on zolpidem vs placebo (*B=3.09, p=0.04, d=0.48*), and evidence for no difference in prior precision between placebo and drug condition in controls (*B=0.25, p=0.86, d=0.07*). There was strong evidence for a difference between controls and patients in the placebo condition (*B=20.4, p<0.01, d=1.12*) such that controls had higher levels of prior precision than patients, but no evidence for or against a difference between controls and patients in the drug condition (*B=0.86, p=0.11, d=0.5*). There was no evidence for or against an interaction between group and condition (*B=1.31*).

The second most important result was that across all participants (patients and controls), there was very strong evidence for a correlation between prior precision and apathy as assessed by AES-Self (*r=-0.46, B>100, p<0.001*), as well as moderate evidence for a correlation between prior precision and AES-Carer (*r=-0.33, B=5.82, p=0.01*) and CamQUAIT-M (*r=-0.29, B=3.07, p=0.02*; see *figure 3*). When separating by diagnostic groups, there remained moderate evidence for a positive correlation between prior precision and the AES-Self in controls (r=-0.37, *B=3.44, p=0.02*), but no evidence for or against a correlation when using the AES-Carer (*r=0.18, B=0.59, p=0.36*) and CamQUAIT-M (*r=0.23, B=0.73, p=0.25*). There was no evidence for or against a correlation between prior precision and apathy in patients (AES-Self: *r=-0.3, B=1.61, p=0.06*; AES-Carer: *r=-0.26, B=0.91, p=0.16*; CamQUAIT-M: *r=-0.23, B=0.77, p=0.21*).

**Figure 3.**
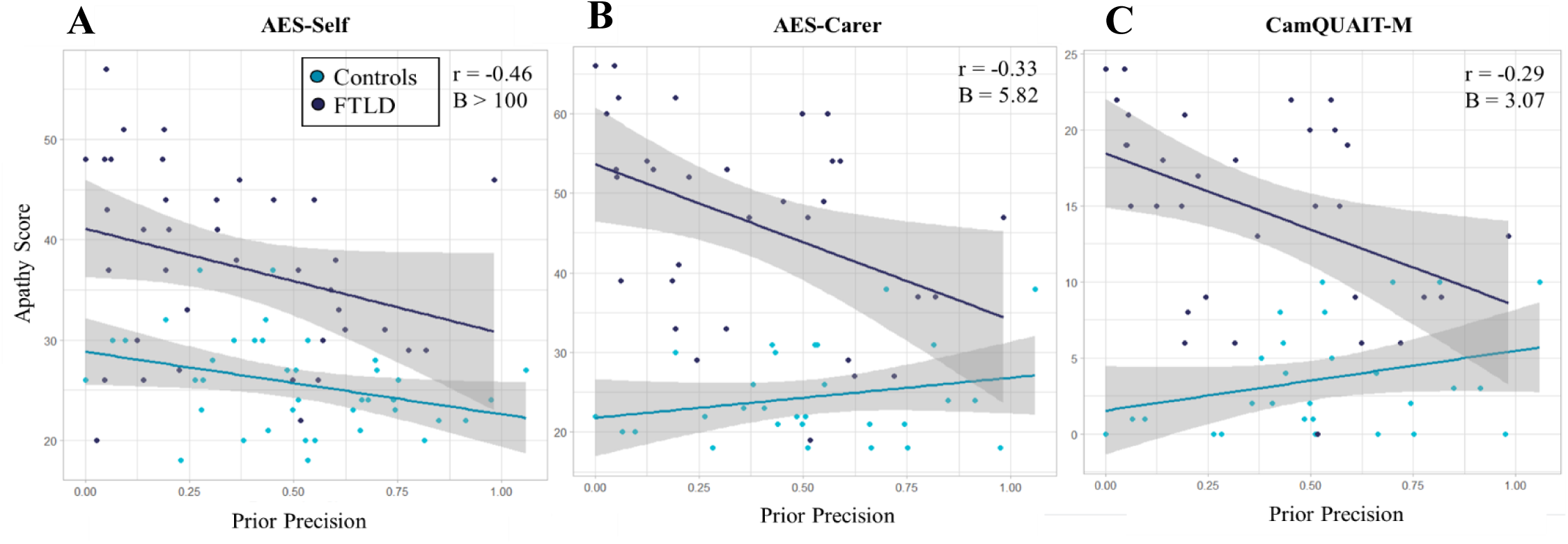
The correlation between apathy and prior precision as assessed by (A) the AES-Self, (B) AES-Carer, and (C) CamQUAIT-M. Reported statistics are for the combined group of controls and patients.

When collapsing across group and condition, there was no evidence for or against a correlation between age or education and prior precision (Age: *r=-0.16, B=0.6, p=0.18,* Education: *r=-0.24, B=2.1, p=0.03*). With separate groups, there was no evidence for or against a correlation between prior precision and age or education in patients (Age: *r=-0.15, B=0.52, p=0.36*, Education: *r=0.008, B=0.36, p=0.96*) or controls (Age: *r=-0.09, B=0.42, p=0.58*, Education: *r=0.2, B=0.65, p=0.23*). Partial correlations were run to explore the impact of including age and education on the relationship between apathy and prior precision but there were no significant effects. There was no evidence for a correlation between the AES-Self and AES-Carer in either controls (*r=0.16, B=0.54, p=0.4*) or patients (*r=0.17, B=0.58, p=0.47*). There was very strong evidence for a correlation between the AES-Carer and CamQUAIT-M in both groups (*r=0.93, B>1000, p<0.001 for both groups*).

### Dynamic Causal Modelling

We compared extrinsic and intrinsic connectivity between controls and individuals with FTLD-associated disorders in both drug and placebo conditions (see *figure 4*). In the placebo condition, there was strong evidence of a reduction in extrinsic connectivity between the premotor and prefrontal cortex in patients compared to controls. In the drug condition, there was evidence that the reciprocal connection from the prefrontal to premotor cortex was increased in individuals with FTLD. In both conditions forward connectivity from the motor to premotor and prefrontal cortex was increased in FTLD versus controls while backward connectivity from the motor to premotor and prefrontal cortex was reduced.

**Figure 4.**
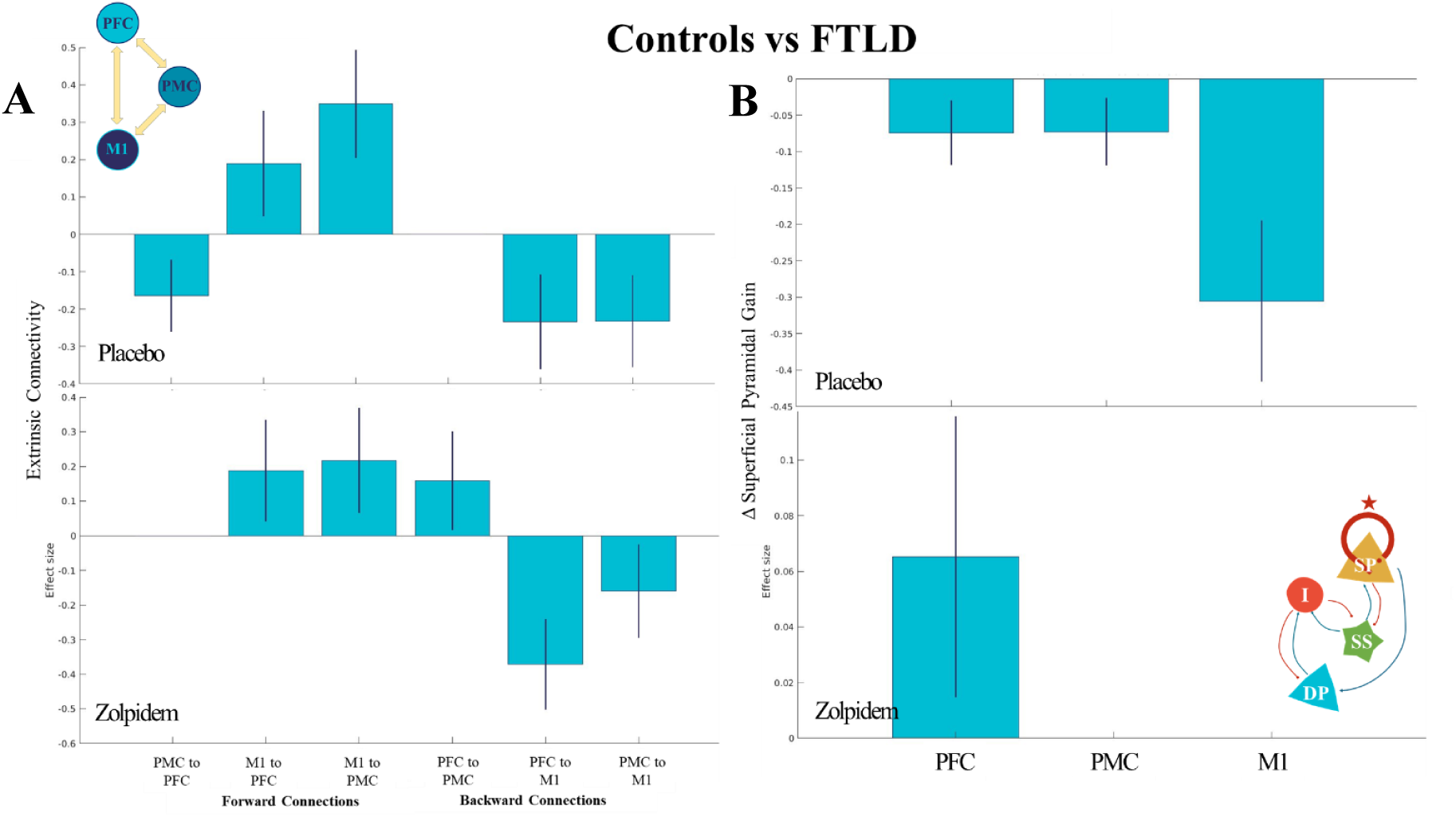
The difference in (A) extrinsic connectivity and (B) superficial pyramidal gain in the placebo and drug conditions. On the y axis, 0 is the mean across patients and controls, positive values indicate parameters that are higher in the patient group than controls, and negative values parameters that are higher in controls than patients.

Regarding the difference in gain on the prefrontal superficial pyramidal neurons, there was strong evidence that gain was reduced in patients compared to controls in the placebo condition but not in the zolpidem condition. Gain on the superficial pyramidal neurons in the premotor and motor cortex was reduced in patients compared to controls in the placebo condition but was not different from controls in the zolpidem condition.

The second model compared drug conditions for controls and patients. There was no evidence of a difference in extrinsic connectivity between drug conditions for controls, but strong evidence of an increase in the connectivity from the premotor to prefrontal cortex in patients on zolpidem compared to placebo *(Pp>0.99)*. In controls, there was strong evidence for a decrease in gain on the superficial pyramidal neurons following GABAergic modulation using zolpidem across all three brain regions. However, there was evidence of an increase in gain in the prefrontal cortex in individuals with FTLD-associated disorders.

Models using only parameters reflecting GABA time constants had higher model evidence than those incorporating only AMPA or NMDA time constants when explaining the difference between conditions in both patients (*∇AMPA=466.09, ∇NMDA=378.89*) and controls (*∇AMPA=197.15, ∇NMDA=160.16*; see *figure 5*). However, GABA time constants were not sensitive to drug effects.

**Figure 5.**
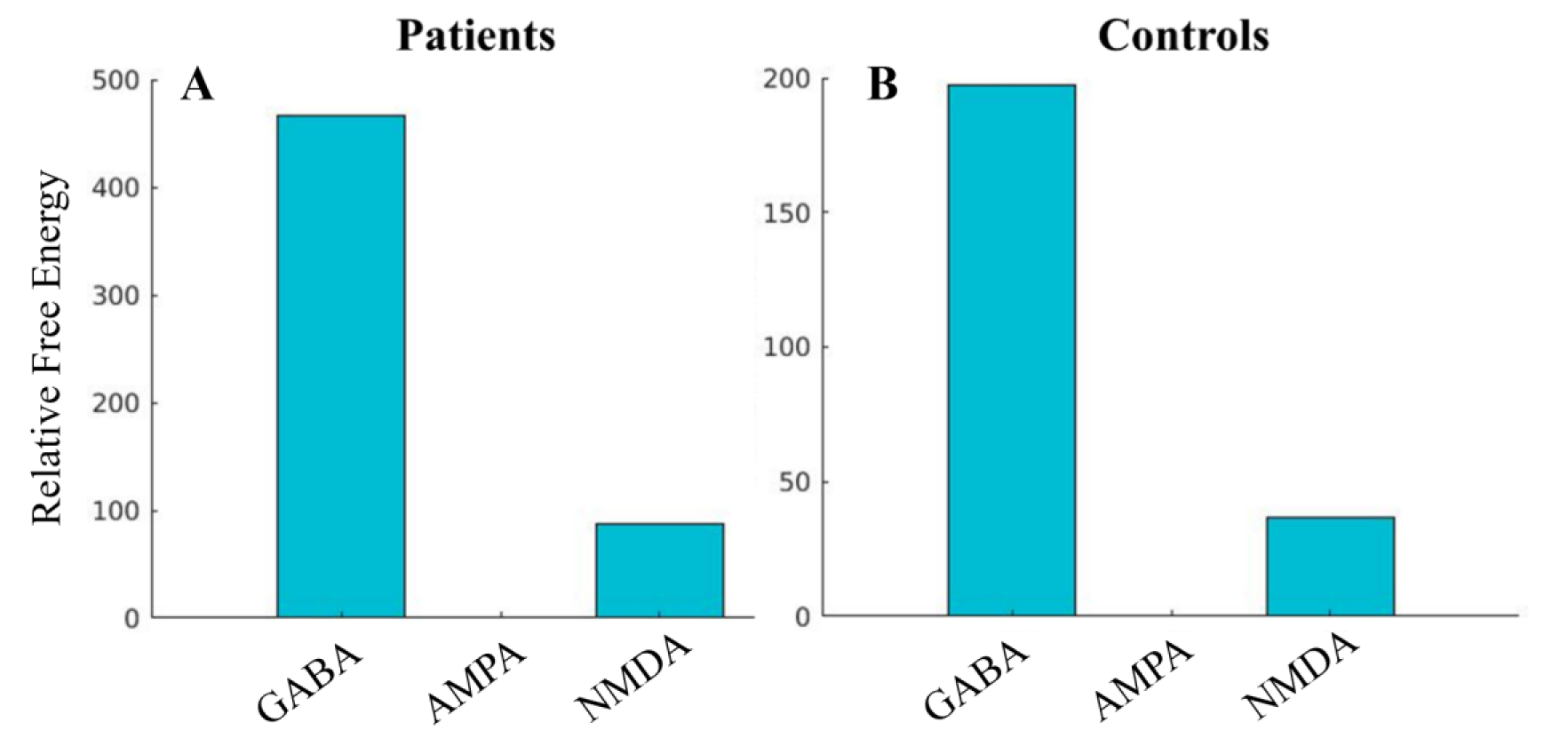
A comparison of free energy for models featuring only GABA, AMPA or NMDA time constants. These are models of the difference between drug and placebo in (A) patients with FTLD and (B) controls

The third set of (PEB) models incorporated individuals’ prior precision as an empirical prior. In the placebo condition, lower prior precision in those with FTLD-associated conditions was associated with higher reciprocal connectivity between the prefrontal and premotor cortex, however this association altered in the drug condition such that lower prior precision was associated with lower reciprocal connectivity between these regions (see *figure 6A*). In the placebo condition, lower prior precision was also associated with higher levels of gain on the prefrontal and premotor superficial pyramidal neurons whereas in the drug condition lower levels of prior precision were associated only with lower gain on the prefrontal superficial pyramidal neurons (see *figure 6B*).

**Figure 6.**
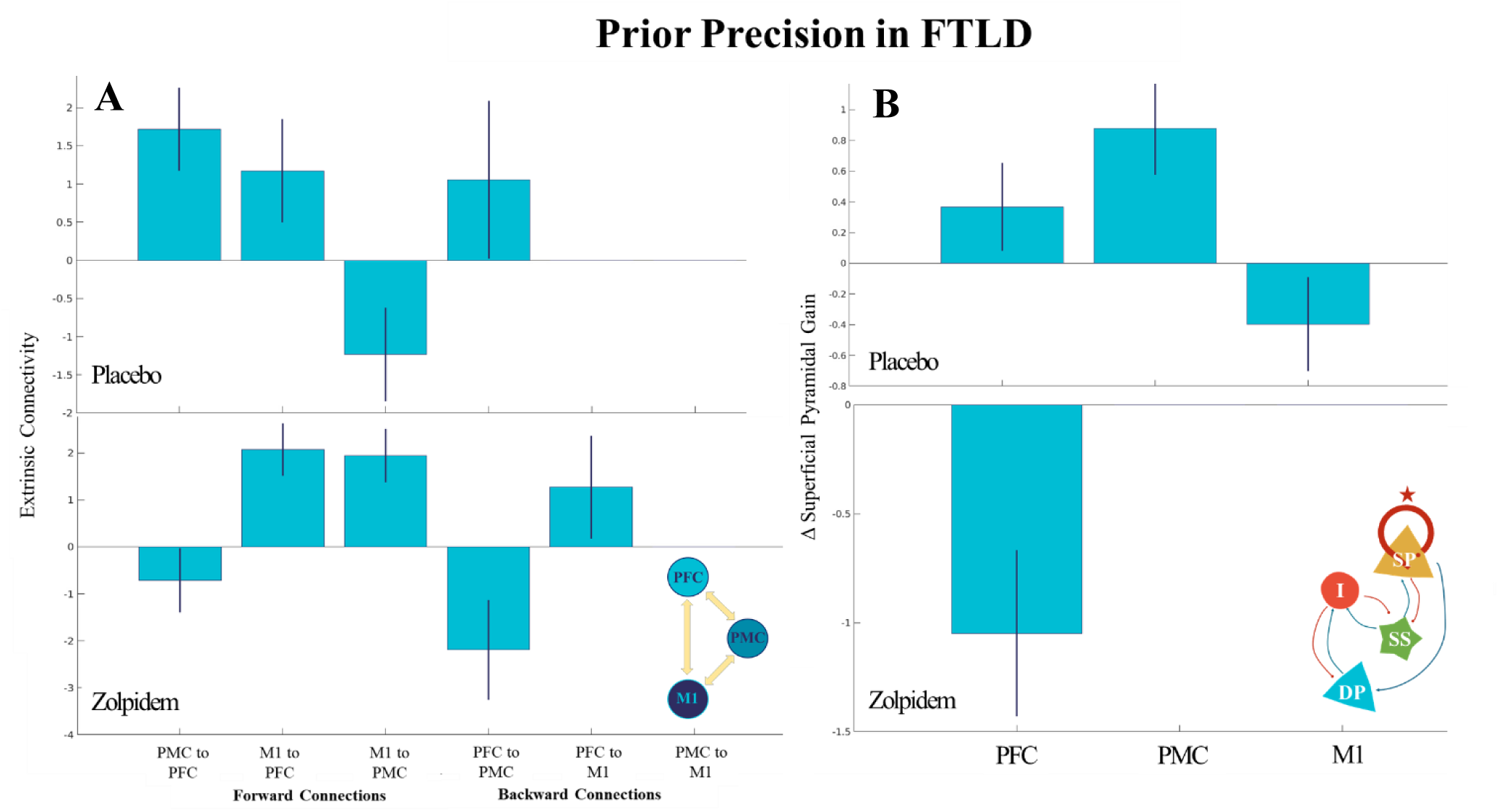
(A) Extrinsic connections and (B) superficial pyramidal gain modulated by prior precision with a posterior probability >0.99. Both extrinsic connectivity and superficial pyramidal gain are shown in placebo and drug conditions in people with FTLD.

## Discussion

There are three key insights from this study. First, we confirm the role of prior precision as a mechanistic explanation for apathy in two disorders associated with frontotemporal lobar degeneration. Second, we show that prior precision can be partially restored with the GABA_A_ agonist zolpidem in people with frontotemporal lobar degeneration. Third, dynamic causal modelling of magnetoencephalography confirmed that the effect of zolpidem may be in part due to changing the postsynaptic gain of prefrontal superficial pyramidal neurons, and subsequent restoration of connectivity in the prefrontal-premotor network. This work highlights the potential for treating apathy by GABAergic restoration in disorders associated with GABA deficits.

As expected, people with bvFTD and PSP differed from controls in apathy on all three qualitative measures (AES-Self, AES-Carer & CamQUAIT-M); and had lower levels of prior precision on the task in the placebo condition. This group difference was abolished on zolpidem, primarily driven by an increase in prior precision in patients but not controls. We also confirmed a correlation between apathy and prior precision. The study was underpowered to test this correlation in the patient group alone (i.e. data lacked sufficient precision for Bayesian inference), though trends were in the expected direction and of similar magnitude in both controls and people with FTLD-associated disorders.

Dynamic causal modelling of neurophysiological observations supported novel mechanistic insights. The reciprocal connectivity between the prefrontal and premotor cortex, and gain on the prefrontal superficial pyramidal neurons, are particularly relevant correlates of apathy and markers of the zolpidem drug effect. Patients had stronger forward connectivity than controls, but weaker backward connectivity from prefrontal cortex. This suggests that behavioural decisions may be dominated by ascending sensory input without the appropriate top-down modulation according to goals and context^62^.

Of particular relevance, the connectivity between the prefrontal and premotor cortex was reduced in patients in the placebo condition but partially restored on zolpidem. In the placebo condition, gain on the superficial pyramidal population was also reduced across all three regions in individuals with FTLD compared to controls. However, this was partially restored on zolpidem, such that prefrontal gain was higher than average in the drug condition. This may be partially driven by a reduction in gain across all three sources in controls on zolpidem, consistent with baseline-dependency by which drug effects are subject to an individual’s baseline.

The dynamic causal modelling of MEG resting-state data also confirmed the associations between prior precision (as the index of apathy) and network connectivity both on and off a GABAergic agonist. In patients on placebo, prior precision was negatively associated with reciprocal connectivity between the prefrontal and premotor regions, but the direction of this relationship reversed in the drug condition such that higher levels of prior precision were associated with higher levels of connectivity. Similarly, there was a negative association between prior precision and gain on the superficial pyramidal neurons in both the prefrontal and premotor cortex when on placebo, but this relationship changed on drug such that higher prior precision was associated with higher prefrontal gain. This mirrors healthy elderly controls who demonstrated a similar association between prior precision and prefrontal gain^9^. Together, these results suggest that prior precision may be mediated by gain on superficial pyramidal neurons, which tunes effective communication across the prefrontal-premotor-motor hierarchy, subject to optimal GABAergic innervation.

There are several limitations to the current study. Firstly, despite the use of Bayesian statistics confirming sufficient precision (c.f. power) in the majority of our analyses, we were unable to draw inferences either way for some comparisons (1/3 < B <3 indicating insufficient precision in the data, analogous to lack of power for frequentist tests). Variants of the ‘Goal Prior Assay’ may improve power in future studies, to quantify the precision of priors on action outcomes; for example, with additional trials or task simplification to encourage meaningful engagement. Incorporating measures which are not reliant on verbal or manual self-report^63^ (e.g. eye tracking) could be an advantage, given the challenges to the interpretation of self-report measures for people with frontotemporal dementia^63^. In addition, the imaging analysis here compares out-of-scanner task performance to in-scanner neurophysiology, albeit on the same study visit. Though we are able to quantify network dynamics and its causes from baseline resting-state analysis, task-based imaging may be used in future studies to further quantify the neural mechanisms at play. There are further limitations relating to the use of zolpidem. Though GABA-A receptors are reduced in individuals with PSP^37^, the evidence is limited and GABAergic deficits in FTLD may be more widespread than can be targeted with zolpidem alone. Moreover, zolpidem acts across the brain, whereas GABAergic deficits are relatively localised in FTLD^38^. This may lead to overdosing of GABA-replete brain regions, resulting in side-effects. Therefore, though this study suggests that GABA agonism may be an effective mechanism to improve prior precision, further research is needed to identify net clinical benefit and examine more selective interventions. Finally, results in people with FTLD may have selection bias (including acceptance bias and completion bias), with more capable individuals completing both the task and resting-state scan. The patient group in the current study therefore represents only a subgroup of the wider patient community, and one with limited diversity.

Future research directions include the extension to other conditions in which apathy is highly prevalent, such as Parkinson’s disease, Alzheimer’s disease^64,65^, depression and schizophrenia^66,67^. It remains to be shown whether prior precision contributes to apathy across all these conditions (noting that it does so in Parkinson’s disease^29^). It is possible that apathy-associated diseases vary in the functional anatomical changes underlying apathy, or the differential contributions of prior precision, effort avoidance and reward sensitivity^68,69^. Exploiting the effect of GABAergic interventions on apathy into clinical trials would also be an important next step, given that the current study was exempted from clinical trials status (in terms of the regulatory framework for clinical trials of interventional medicinal products) as we did not investigate the impact of zolpidem on clinical endpoints. Baseline dependency also remains to be tested as a treatment-decision criterion, implementing a precision medicine approach to treat according to an individual’s GABAergic deficit rather than clinical diagnostic group assignment.

In conclusion, we show that apathy in two disorders associated with frontotemporal lobar degeneration is underpinned by the loss of prior precision on action outcomes and a subsequent failure of active inference. This loss of precision is accompanied by a reduction in gain on the prefrontal superficial pyramidal neurons and subsequent functional disconnections across the prefrontal-motor decision-making hierarchy. The GABAA-agonist zolpidem partially restored prior precision, and frontal network connectivity, opening potential new approaches to treat apathy.

## Data availability

Data supporting the work in this publication is available on request from the corresponding author. Fully anonymized, post-processing MEG data is available on request. Clinical metadata are also available on request but will likely require a data transfer agreement to comply with confidentiality and consent.

