## Supplemental Material 1 for "GABAergic regulation of action-outcome priors in conditions associated with frontotemporal degeneration"

### Supplementary Material 1

Demographic and clinical characteristics of individuals with FTLD, split into 'responders' and 'non-responders' to the 'Goal Prior Assay Task'. Participants with relevant data missing were excluded at the level of individual analyses.

| | Responders (N=23) | Non-Responders (N=17) | Comparison |
|---|---|---|---|
| Age | 68.05 (7.82) | 71.1 (9.28) | B = 0.5 (p = 0.38) |
| Gender (M:F) | 16:6 | 8:6 | B = 0.64 (p = 0.55) |
| Education | 12.74 (3.29) | 12.08 (2.06) | B = 0.39 (p = 0.46) |
| Diagnosis | 11:12 | 10:7 | B = 0.47 (p = 0.71) |
| *ACER* | *83.05 (11.1)* | *61.92 (20.45)* | *B = 51.7 (p < 0.01)* |
| CBI | 69.33 (44.56) | 67.61 (25.91) | B = 0.34 (p = 0.89) |
| AES-Self | 37.13 (9.79) | 42.83 (12.31) | B = 0.79 (p = 0.18) |
| AES-Carer | 46.7 (12.67) | 51.91 (9.55) | B = 0.59 (p = 0.21) |
| CamQUAIT | 14.85 (6.54) | 19 (6.43) | B = 0.96 (p = 0.11) |
